# Adolescent Voices on Global Racisms: Lived Experiences, Health Impacts and Demands Across Six Countries

**DOI:** 10.64898/2026.09.02.26361744

**Authors:** Priscila de Morais Sato, Sonora English, Jenevieve Mannell, Sorcha Ni Chobhthaigh, Hanui Choi, María Calderón, Heizal Patricia Nagginda, Patrick Hans Mulindwa, Aloka Weerasekara, Seung-Sup Kim, Kusala Wettasinghe, Ananda Galappatti, Ran Li, Mita Huq, Zey Binboga, Caroline Parker, Delan Devakumar

## Abstract

**Introduction:** Although the mechanisms of racism may be globally consistent, both these mechanisms and their context-specific expressions require further investigation. Comparing perceptions and experiences of minoritised adolescents across countries helps reveal how racism manifests in different contexts. This multi-country study investigates how adolescents understand, experience, and demand action against the racisms that shape their health and lives.

**Methods:** We conducted semi-structured interviews with 93 adolescents from minoritised groups across Brazil, Peru, South Korea, Sri Lanka, Uganda, and the UK, using culturally adapted vignettes. Using inductive thematic analysis, country-based and external coders applied a co-developed cross-country codebook, achieving high inter-rater reliability, and identified seven key themes.

**Results:** Participants’ narratives showed how appearance, language, and socioeconomic status functioned as social markers through which racism reproduced inequities in education, healthcare, and economic opportunity. Across contexts, racism imposed psychological strain, through social exclusion combined with pressure to succeed. This affected adolescent health, contributing to emotional distress, low self-esteem, and uncertainty, while shaping help-seeking, institutional trust, and exposure to material conditions that worsened physical health. Experiences varied: ethnic diversity in the UK offered perceived protection from interpersonal discrimination; economic security in South Korea helped render a minoritised identity an asset; while spatial segregation in Brazil, Peru, Sri Lanka, and Uganda reduced external threat but intensified structural disadvantage. Adolescents called for stronger support networks, anti-racist education, accountability, improved opportunities, and broader justice in workplaces and online spaces.

**Conclusion:** Our findings underscore that addressing racism’s impacts on adolescent health requires a health justice approach. This must recognise racism as a public health crisis, confront racialised structural inequities, promote resource access, and engage adolescents in policy development.

## INTRODUCTION

Racism is increasingly recognised as a critical public health issue and a fundamental driver of global health inequities. Its effects extend beyond physical health, shaping the social determinants that influence life opportunities and outcomes.^1^ Among adolescents, these consequences are particularly profound. Evidence links racism, xenophobia, and other forms of discrimination to mental distress, emotional and behavioural difficulties, low self-esteem, reduced academic engagement, and poorer physical health for adolescents.^2^ Adolescence is a formative stage of development characterised by rapid biological and psychosocial change.

During this period, young people construct their identities, values, and sense of belonging, making them particularly sensitive to exclusion and discrimination that can have lasting impacts on well-being and self-perception.^3,4^

Racism operates through interconnected structures of separation and power: (1) structural, rooted in historical systems that sustain inequity; (2) institutional, through policies that restrict access to resources and opportunity; (3) spatial, by concentrating minoritised groups in environments that undermine health; (4) community, where discrimination coexists with resilience; and (5) individual, through interpersonal and internalised forms of discrimination.^1^ Although the mechanisms of racism are globally consistent, their expressions are shaped by distinct social, cultural, and historical contexts. The concept of *global racisms* captures the interconnected forms of racism rooted in colonial legacies and sustained by contemporary political, economic, and social processes. Global racisms constitute a global health problem, shaping exposure to risk, access to care, and the distribution of health itself, while legitimising inequity and producing preventable disparities in well-being and life expectancy.^5^

Existing research on racism and adolescent health has primarily focused on behavioural and mental health outcomes, which constitute most of the evidence base, while comparatively fewer studies have examined physical health outcomes or healthcare experiences.^1^ Within this literature, racism is conceptualised as a chronic and cumulative stressor, with evidence linking exposure to psychological distress, behavioural responses, and emerging physiological processes.^4^ Intergenerational dynamics have also been documented, with parental experiences of discrimination associated with child internalizing problems through pathways such as parental mental health and parenting practices.^2^ Empirical work has largely operationalised racism at the interpersonal level, with more limited attention to structural and institutional dimensions, including within health and social systems.^3^

Despite these contributions, the scope of the evidence remains uneven. Studies are concentrated in the United States and predominantly focus on African American populations, with limited attention to other minoritised groups and to variation across geopolitical contexts.^1–3^ In addition, many studies rely on parental reports or adult-derived measures, with fewer drawing directly on adolescents’ own accounts of their experiences.^2^ There is also limited qualitative, youth-centred research examining how adolescents interpret and make sense of racism in their everyday lives and how they identify priorities for addressing its health impacts, particularly across diverse global settings.

Race and related concepts, such as ethnicity, colour, and caste, are socially produced relations of power rather than biological facts. Systems for classifying human difference differ widely among the study countries, reflecting distinct historical, social and political trajectories. The United Kingdom census uses an ethnic group framework, Brazil asks directly about race, Peru, Sri Lanka, and Uganda refer to ethnicity or tribe, and South Korea does not formally classify by race or ethnicity. These variations show that race is a fluid social construct shaped by specific cultural and institutional contexts.^6^ Comparing perceptions and experiences of minoritised adolescents across countries offers critical insights into how global racisms manifest locally, confirming shared mechanisms and revealing context-specific expressions. Such understanding can inform adolescent-led, contextually grounded, and temporally relevant responses. Through qualitative research with minoritised adolescents across six countries and four continents, we explored how racism is understood and experienced and identified young people’s priorities for addressing its health impacts. Moving beyond documenting harm, we examine cross-contextual patterns and highlight youth-driven solutions.

## METHODS

### Participants and Recruitment

We conducted semi-structured interviews with adolescents aged 15-17 years old from minoritised groups in Brazil, Peru, South Korea, Sri Lanka, Uganda and the United Kingdom (Figure 1). Minoritised groups are not necessarily numerical minorities; rather, they are constituted through social, political, and historical processes that produce unequal power relations.^7^ While minorisation can intersect with multiple social categories, this study focuses on minorisation as a product of racialisation, including how racialised meanings are shaped through ethnicity, indigeneity, and migration-related belonging. Country selection drew on existing research partnerships while also aiming to capture diverse social, political, and historical contexts in which racism and its health impacts manifest.

**Figure 1.**
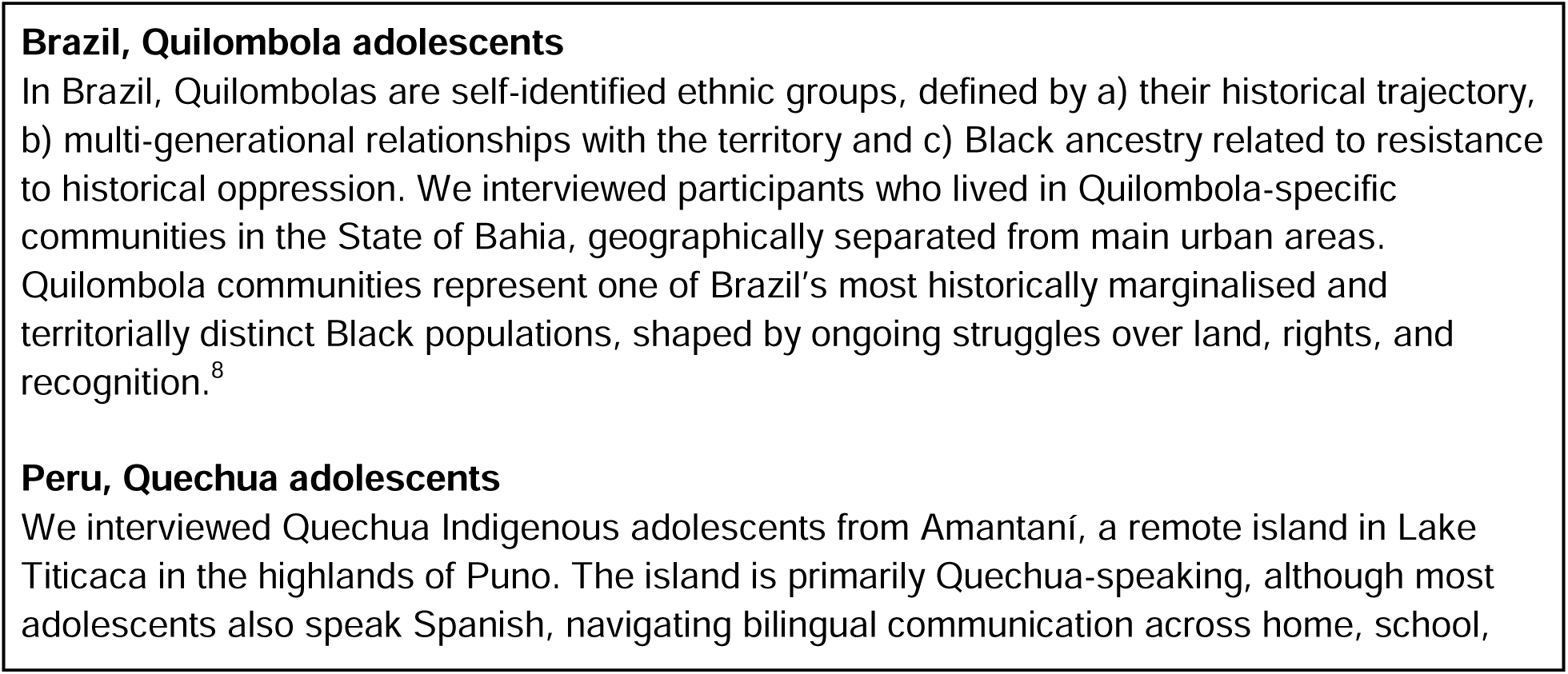
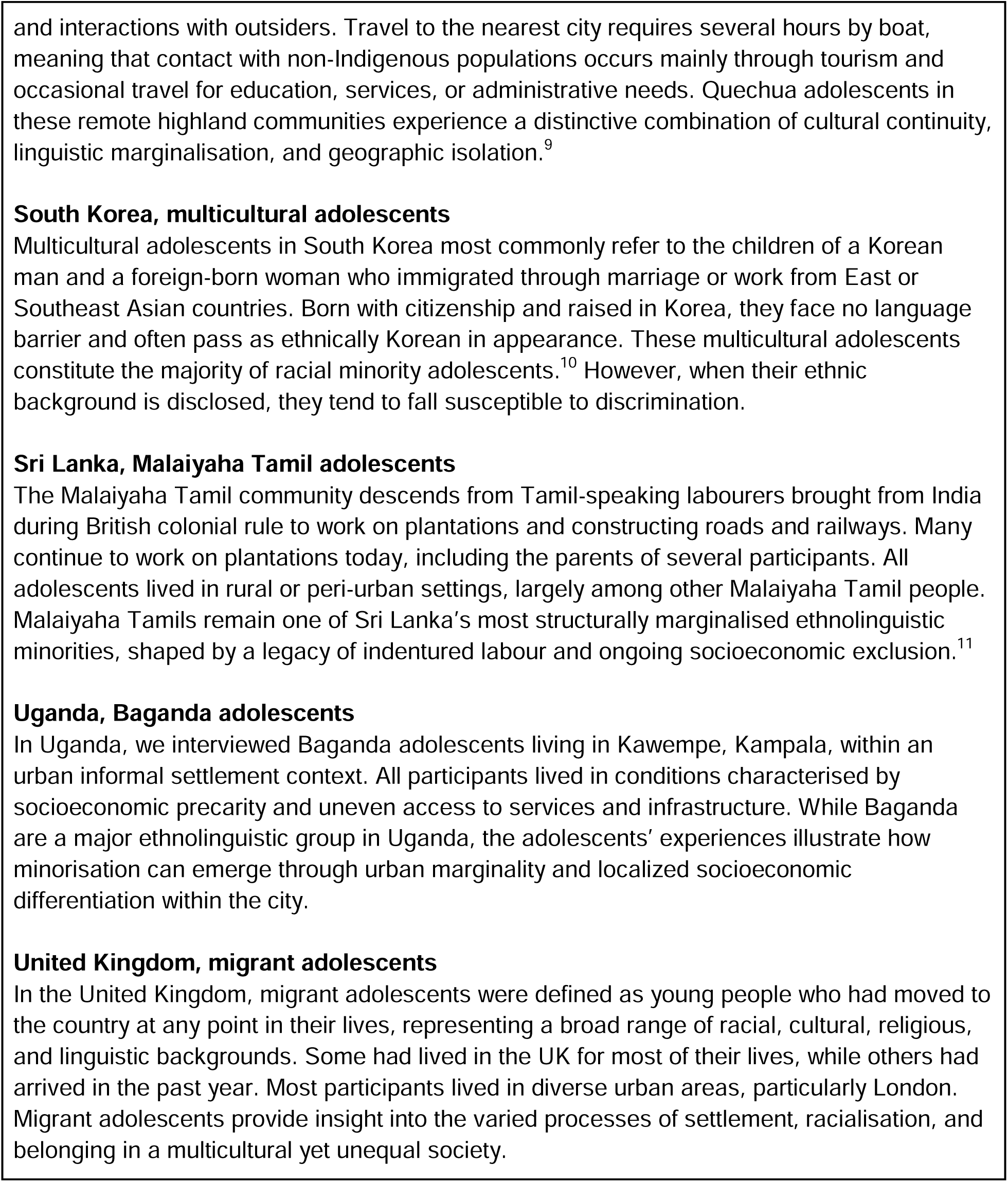
Participant Characteristics and Context Across Study Settings.

The project’s purpose was explained and informed consent obtained prior to interview. Interviews in each country were conducted until data saturation was achieved.^12^ In Brazil, Peru, Uganda, South Korea, and Sri Lanka, parental consent and participant assent were obtained, while in the United Kingdom, participants provided their own informed consent. In the UK and South Korea participants were compensated with vouchers, and in Peru and Uganda with school materials. They were not compensated in other countries. Differences in consent and compensation procedures reflected variations in local norms and regulatory requirements across sites. Table 1 summarises information about participants and recruitment.

**Table 1:** Sample, recruitment and interviews.

| Country | Participants | Recruitment | Data generation | Interview language |
| --- | --- | --- | --- | --- |
| Brazil | Quilombola youth (n=19) | In partnership with community associations | In person and online, by a researcher with previous work with the community | In Portuguese |
| Peru | Indigenous youth (n=20) | In partnership with a local global health institution | In person, by researchers with previous work with the community | In Spanish |
| South Korea | Multicultural youth (n=12) | In partnership with migrant workers organisations | In person and online, by researchers | In Korean |
| Sri Lanka | Malaiyaha Tamil youth (n=19) | In partnership with a locally based health policy and research institution | In person, by researchers with previous work with the youth | In Tamil |
| Uganda | Baganda youth (n=11) | In partnership with a grassroots organization | In person, by volunteers with previous work with the community | In Luganda and English, according to participant preferences |
| United Kingdom | Migrant youth (n=12) | In partnership with a community-based organisation | In person and online, by researchers with previous work with the youth | In English or Spanish, according to participant preferences |

Ethical approval was obtained from all participating institutions: the Ethics Committee of the School of Nutrition of the Federal University of Bahia, Brazil (No. 6,598,386); the Institutional Bioethics Committee of Vía Libre, Peru (No. 10706); the Institutional Bioethics Committee, South Korea (IRB No. 2404/003-014); the Institutional Review Board of Institute for Health Policy, Sri Lanka (No. IRB/2024/037); the Ethics Board of Uganda Christian University, Uganda (UCUREC-2024-937); and the University College London Research Ethics Board, UK (No. 27091/002).

### Data Generation

Interviews were conducted one-to-one in a local language by local trained interviewers (Table 1). A semi-structured interview guide comprising a vignette followed by open-ended questions was used across all countries. Vignettes are a valuable tool for exploring sensitive topics,^13^ and were adapted in each country to ensure cultural relevance. Each presented a short story about two characters, one from a minoritised group and one from a non-minoritised group, with differing experiences in education that alluded to institutional and structural racism. Educational institutions were chosen as 15–17-year-olds are most likely to interact with them. By alluding to structural and institutional racism the vignettes encouraged discussion of racism beyond interpersonal examples. The interview guide was reviewed by minoritised and non-minoritised youth advisors in the UK and Romania to enhance its content, clarity, and relevance. Youth advisors were recruited through *The Lancet Commission on Racism and Child Health*, in partnership with Save the Children.

Participants were asked what made the characters different, how these differences impacted their lives, and how this could make them feel. They were then asked about their own experiences of discrimination or those of people close to them, and what changes would make the world fairer for adolescents and children. Finally, participants were asked about any other thoughts they would like to share. Probing questions were used throughout to elicit detailed responses. Interviews lasted 32 minutes on average.

Reflecting language, cultural, and contextual differences, different language was used across sites to refer to racism. For example, “discrimination” was more commonly used as an accessible term for adolescents in some contexts, while “racism” was used less frequently and in some settings appeared less readily used in interview discussions. With consent, interviews were recorded, transcribed verbatim and translated to English for analysis.

### Data Analysis

We conducted an inductive thematic analysis in four stages:^14^

(a) *Pre-analysis:* Two researchers (PMS, SE) reviewed all transcriptions multiple times to familiarise themselves with the data and identify inductive, exploratory codes for each country.

(b) *Material exploration:* Exploratory codes were grouped by similarity to form themes and subthemes which were discussed with researchers from all countries (AG, AW, CP, DD, HC, HM, HPN, JM, KW, MC, MH, RL, SNC, S-SK, ZB).^15^ A codebook was co-developed and refined with researchers from all countries, including for each subtheme: short and detailed descriptions, inclusion/exclusion criteria, and typical, atypical, and ‘close but no’ examples.

(c) *Data treatment:* Two coders, one country-based (HC, HM, HPN, KW, MC, PMS, SNC) and one external (PMS or SE), independently applied the codebook to two interviews per country using MAXQDA. Inter-rater reliability was assessed for each country using Cohen’s Kappa coefficients. Kappa values below 0.8 triggered discussions to resolve discrepancies and refine understanding before re-coding. Once Kappa reached >0.8 across all subthemes, a researcher familiar with each country’s data applied the codebook to the full dataset. A single researcher (PMS) then reviewed all coded interviews to ensure consistency and accuracy across the dataset.

(d) *Inference and interpretation:* We compared themes and subthemes across countries by examining how racism operated at structural, institutional, and individual levels, and how these levels interacted. This approach enabled the identification of variations in expression and shared mechanisms through which racism impacts health.

## RESULTS

Ninety-three adolescents participated in the research. Inductive thematic analysis generated 32 subthemes, organised into seven themes: perceptions of difference, negotiating racism, forms and contexts of discrimination and racism, health consequences of racism, approaches to navigating racism, existing forms of support and protection, and demands to make the world a fairer place. The sections below present the themes and subthemes, highlighting cross-country differences, contextual nuances, and illustrative examples. Detailed definitions of each subtheme and additional country-specific quotations are provided in the <u>Appendix</u>.

### Minoritised adolescents’ perceptions and understandings of differentiation and racism

Perceptions of difference referred to how participants distinguished adolescents from minoritised and non-minoritised groups. These distinctions were described in relation to cultural background, including differences in traditions, customs, religion, habits, clothing, and broader notions of “culture”; appearance, particularly skin tone, hair texture, and facial features; language, including accent, languages spoken, and proficiency; location, referring to where minoritised adolescents had grown up or currently lived; and access to economic and educational resources. These differences were not necessarily perceived as negative, despite participants noting the role of differences in separation and discrimination.

Participants in Brazil, Peru, South Korea and the UK named cultural background as the main differentiation between minoritised and non-minoritised adolescents, followed by appearance in Brazil, Peru, Sri Lanka, and the UK. In South Korea, where non-Korean parents were typically from neighbouring Asian countries, participants did not consider physical appearance a significant marker of difference. Location was mentioned by participants from Brazil, Peru, Sri Lanka and Uganda and established worse educational and economic resources, lower status, and lower access to infrastructure and services.

Participants’ responses to racism ranged from realisation and active critique to normalisation and internalisation. Realisation of racism was mentioned in all countries, with more frequent mentions in Brazil, Sri Lanka and the UK: “Like, in society we live in people with lighter skin tones, like in every aspect in life, are treated better than people with darker skin tones” (Girl, UK). Many participants simultaneously acknowledged and critiqued racism while also normalising it. In South Korea and the UK, the minimisation of racism was linked to perceiving discrimination as an individualised phenomenon. For instance, verbal attacks were often interpreted as jokes as a coping strategy to mitigate psychological discomfort. For participants in Sri Lanka, minimisation of racism was reflected in a more accepting or matter-of-fact stance toward discrimination, expressed through sentiments such as: “I take it as it is by thinking ‘This is how life is’, ‘That is how they will be’ and I won’t let it bother me” (Girl, Sri Lanka). Normalisation of racism was not explicitly mentioned in Brazil, Peru and Uganda. Internalisation of racism was reported by participants in Brazil, Peru, South Korea, and the UK, shaping identity, self-worth, and social participation. In Brazil, it appeared as Quilombola identity denial, while in the UK it manifested as the internal acceptance of harmful stereotypes or marginalisation: “So people can just base you off that and think you’re dumb… you can get lower grades, that actually happens” (Boy, UK). In Peru, economic inequality intersected with internalised social hierarchies, influencing how adolescents perceived their own agency. In South Korea, societal attitudes toward multicultural adolescents were described as fostering internal conflict and self-doubt. By contrast, participants in Sri Lanka and Uganda did not explicitly describe internalisation.

### Global patterns of racism and health consequences

Forms of discrimination differed across countries. At the structural level, lack of access to resources and infrastructure was most relevant in Brazil, Peru, Sri Lanka and Uganda: “[minoritised character from the vignette] might eat poorly because his family can’t afford good food. Living in a slum with poor sanitation also increases the risk of diseases. [Non-minoritised character from the vignette]’s family can afford better food and clothing, and they can quickly get treatment if anyone falls sick (Girl, Uganda). Participants from Brazil, Peru, Sri Lanka, Uganda, and the UK reported worse future opportunities and conditions, which was compounded by limited access to resources in each setting. Discrimination was most frequently mentioned at the institutional level, particularly regarding worse treatment in educational institutions. Lack of availability and accessibility to good education took different forms across contexts. In Brazil, Peru, Sri Lanka, and Uganda, spatial segregation meant that minoritised communities often lacked access to affordable, well-equipped schools that mediates ability to qualify for university entrance. In the UK, language barriers were reported as the main obstacle to education, particularly for more recent arrivals. Within health systems and institutions, participants reported lack of availability and accessibility to healthcare and medicine, including the poor quality and unaffordability of healthcare in Brazil, Peru, Sri Lanka, and Uganda, as well as language barriers in the UK. This was exacerbated by worse treatment in health institutions, through healthcare professionals restricting access for minoritised individuals and through patients avoiding services due to prior interpersonal or familial maltreatment. “When a person from the city arrives, I think they [health workers] will prioritise them, the White person, rather than the Quilombola person, who is Black and with mud in their shoes” (Boy, Brazil).

At the interpersonal level, verbal and written insults, attacks, and discriminatory gazes were mentioned in Brazil, Peru, South Korea, Sri Lanka and the UK. At times, adolescents described not being looked at and effectively being ignored in schools, on the streets, and in other public spaces, reinforcing feelings of invisibility. In Brazil, South Korea, Sri Lanka and the UK, participants reported assumptions of wrongdoing and unequal disciplinary measures in schools and commercial spaces, being unfairly suspected, monitored, or reprimanded. These incidents were most experienced by boys, though girls were also targeted. Even when girls were not directly involved, they were negatively affected by fear or concern for their family members or friends.

> “There was a confusion here, and he was in the middle, but he didn’t do anything, it wasn’t the wrong one. But because he was Black, dark-skinned, he ended up being arrested. I think he spent a night or two there in prison […] because he was mistaken for someone else. Discrimination is a sad thing. Unfortunately, on a daily basis, it is increasing” (Girl, Brazil).

Health consequences of racism were mainly related to poor emotional wellbeing and mental health in all countries. In Brazil, Peru, and Sri Lanka, mental distress was associated with experiences of discrimination and perceptions of limited opportunities: “there are also people who do not treat everyone equally, and they bully, which can affect [us] emotionally” (Girl, Peru). In Peru and Uganda, perceived disparities in material conditions also contributed to feelings of sadness and poor wellbeing: “Sometimes, financial worries can make a person feel bad about how they are going to continue their studies or achieve their goals” (Girl, Peru). Sri Lankan participants frequently emphasised a strong sense of injustice in how they were treated and the limited opportunities they had as a cause of mental distress. In the UK, emotional difficulties stemmed from cultural barriers to socialising and from being judged or ridiculed, which led to insecurity. Some participants from South Korea associated loneliness and stress with poor mental health, describing difficulties opening up and stress tied to the pressure to succeed and uncertainty of achieving future goals.

> “From my own experience, I think there’s definitely a lot of pressure. Like, when you have a dream, it’s exciting, but at the same time, it feels so far from where I am now. I sometimes wonder if I even deserve to get there. And just thinking about how hard I have to work—it’s just kind of overwhelming” (Boy, South Korea)

Perceived physical impacts were mainly related to poor nutrition in Uganda, Sri Lanka, and the UK due to lack of access to sufficient and nutritious foods: “If these people only get two meals or one meal a day, and even that is not proper food, then it will affect their health. Things like gastric issues and many other problems will come up” (Boy, Sri Lanka). Health problems linked to poor sanitation and exposure to pollution were reported in Sri Lanka and Uganda. “If there is a person who is sick in the next house, there is a chance of them getting the sickness. Houses in the plantation have more chances of them having an animal like a goat or cow. Since there won’t be proper care for it, there is a chance they will get infections” (Boy, Sri Lanka).

Participants in Brazil and the UK reported physical harm associated with aggression related to racism. Physical impacts of racism were not mentioned in South Korea.

Most behavioural responses to discrimination were similar across all countries, including working harder, distracting themselves and self-isolating and avoiding places and situations. Confronting the perpetrator was mentioned by few participants, mainly from South Korea, Sri Lanka and the UK.

> “They will say stuff and I shouldn’t be that bothered about it anyway but genuinely, if I see someone being extremely rude to my siblings or like my friend, or any random person of colour, honestly I think I would react to it because I know what it feels like to be that person that’s just there” (Girl, UK)

### Minoritised adolescents’ priorities for action to address the health effects of racism

Adolescents across contexts described their priorities for protecting them from and combatting global racisms, including: buffers and protective mechanisms, and demands to make the world a fairer place. Buffers and protective mechanisms for adolescents were primarily situated at the individual level in Peru, Sri Lanka, and Uganda, where support from family and friends (in Peru and Uganda) and from minoritised teachers (in Sri Lanka) played a central role. Within institutions and systems, adolescents in Brazil, South Korea, and the United Kingdom highlighted measures such as racial and cultural education programs. Affirmative action policies were discussed in Brazil, and South Korea. In the United Kingdom, adolescents also emphasised diversity-oriented policies in hiring and healthcare. Participants felt protected from interpersonal discrimination within communities and local environments, when the community was diverse within an otherwise homogeneous, majority-dominated context (as in the UK) or was composed primarily of a minoritised group (as in Brazil, Peru and Sri Lanka). Korean participants from socioeconomically stable backgrounds occasionally viewed their multicultural identity as an advantage, particularly when proficiency in languages such as English or Chinese supported their educational and career prospects.

Demands to make the world a fairer place were described at different levels. At the individual level, adolescents in Peru and Sri Lanka emphasised the need for equal treatment and support from family and friends. At the institutional level, young people in Brazil, South Korea, and the United Kingdom called for anti-racist education and for rules and consequences to address racism: “Like, teachers just make them do a few hours of community service at school and that’s it. I think maybe the punishments should be stricter — like, transferring them or even expelling them. I just feel like it needs to be taken more seriously” (Boy, South Korea). Across all countries, participants highlighted the importance of better education and job opportunities as essential for fairness. At the structural level, local government officials were seen as mainly responsible for ensuring equity through policies that promote fairness, particularly in the workplace and online spaces: “I think the government needs to ensure fairness, but leaders should guide people to treat others with love and respect (Boy, Uganda).

## DISCUSSION

This study is the first to examine adolescents’ perceptions of racism across different national contexts, its health impacts and their expectations for addressing its health impacts. In Brazil, Peru, Uganda, and Sri Lanka, discrimination based on race and ethnicity intersected with economic disadvantage to intensify exclusion, which reflected in verbal abuse and segregation within under-resourced schools and health services. In the UK, physical appearance, language, and accent were tied to classroom exclusion and racialised surveillance in public spaces. In South Korea, where the myth of ethnic homogeneity and the concept of ’pure Koreans’ are deeply ingrained^16^, multicultural adolescents can be particularly vulnerable to identity confusion and psychological insecurity. Figure 2 presents adolescents’ perceptions of racism at different levels across countries and its perceived impact on adolescent health.

**Figure 2.**
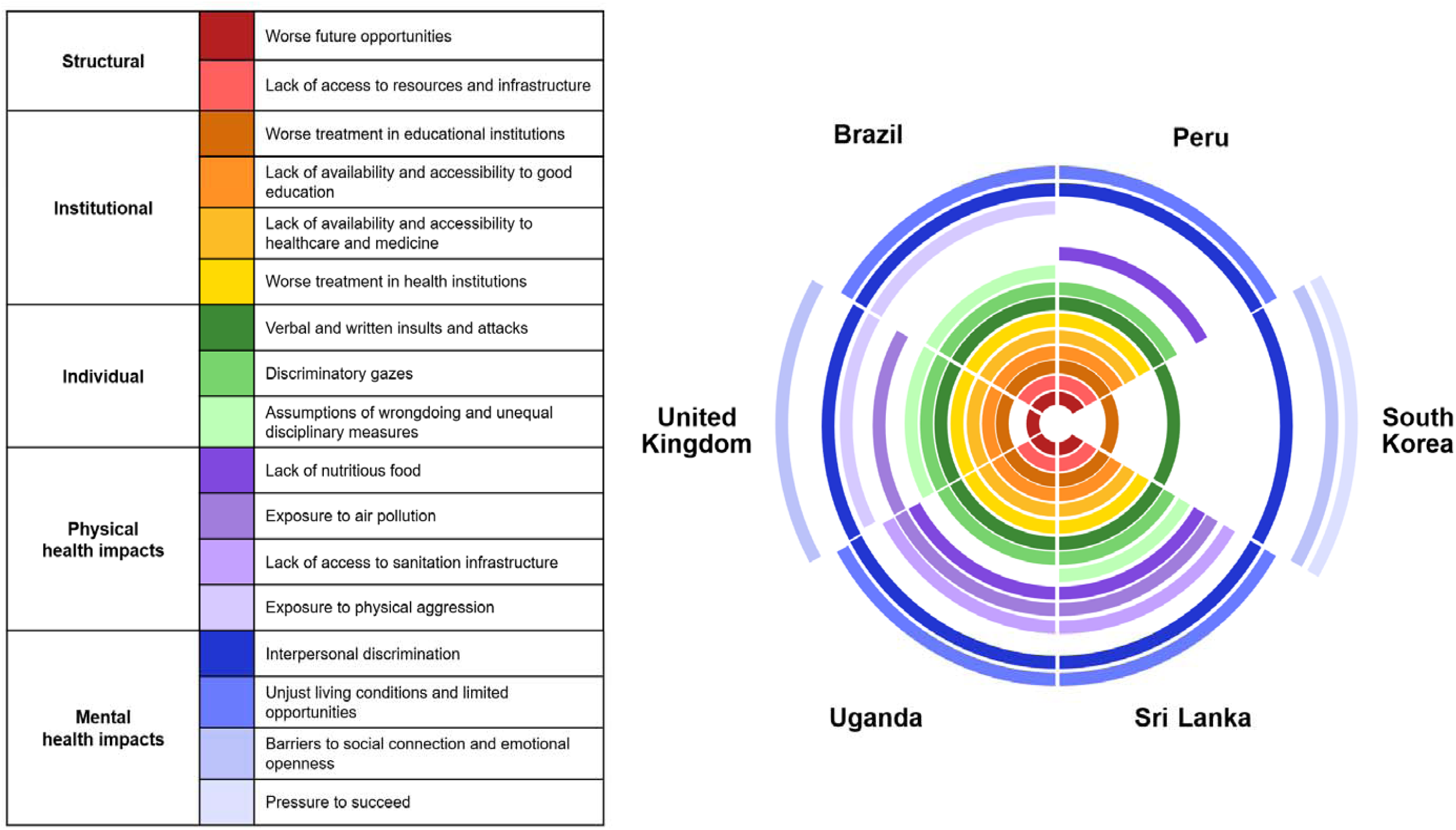
Perceived forms of racism and health impacts among adolescents across countries.

Importantly, the differences described across countries primarily reflect how racism is recognised and interpreted by participants within each context, and only indirectly speak to how it is manifested. The absence of reported structural forms of racism in South Korea, for example, should not be taken as evidence of their nonexistence, but may instead indicate that such forms were not readily identified by participants. Across all countries, different factors produced similar patterns of social exclusion, manifested through bullying and isolation, contributing to psychological distress and adverse health outcomes among adolescents

### Global Racisms

Our findings support the concept of global racisms by documenting related forms of discrimination across countries and identifying a shared pattern in which discrimination is produced through socially and historically embedded markers of difference linked to unequal power relations. These processes contribute to a cyclical dynamic in which discrimination reinforces existing inequalities, which in turn generate conditions for further discrimination. At the same time, the diversity of experiences described across contexts, shaped by intersecting social positions, highlights the varied ways in which these processes are lived and negotiated. Space and location played an important role in differentiating people and shaping forms of racism. While identity was closely tied to where adolescents or their parents came from, spatial segregation varied across settings and influenced how discrimination was experienced. Our findings show that the effects of spatial concentration and segregation vary depending on local community structures and social composition. In some contexts, like Brazil, Peru, Sri Lanka and Uganda, such concentration can function as a protective environment against everyday racism, while movement into majority-dominated spaces may increase exposure to discrimination. In the UK, participants who grew up in more diverse urban environments reported fewer experiences of interpersonal discrimination in neighbourhoods characterised by greater diversity compared to less diverse majority-dominated areas. This aligns with evidence suggesting that sustained, everyday intergroup contact under relatively equal conditions can reduce perceived discrimination.^17,18^ Across contexts, spatial organisation also shaped access to resources and opportunities, with more segregated or marginalised areas often characterised by reduced availability of services and infrastructure, while majority-dominated or more central areas offered greater availability of services, though not always equitable access. In South Korea, while formal residential segregation is limited, many multicultural adolescents are concentrated in less urban areas with comparatively fewer educational, cultural, and social resources than urban centres.

### Health Impacts of Global Racisms

A primary finding of this study is that mental health emerged as the health dimension most prominently described as affected by racism in adolescents’ accounts. This finding is consistent with a scoping review from Brazil, which identified associations between interpersonal racism, and depression and low self-esteem among young people, while also highlighting evidence that racism interacts with socioeconomic disadvantage to increase vulnerability to infectious diseases and poor oral health in this population.^18^ However, our findings extend this literature by showing that, across diverse national contexts, adolescents consistently described psychological wellbeing as the aspect of health most immediately affected by racism.

Participants described how racism shaped identity, self-worth, and social participation through processes of internalisation, although these experiences took different forms across countries, the underlying process was similar. These findings are consistent with studies describing internalised racism as the incorporation of negative social beliefs about one’s own racial group, contributing to chronic stress, diminished self-esteem, and negative self-evaluation.^19^ Previous research has also linked prolonged exposure to racism with more severe psychological outcomes, including hypervigilance, feelings of persecution, and subclinical psychotic experiences.^20^ However, participants did not simply reproduce these beliefs. Adolescents across all countries recognised racism as part of their daily lives and frequently criticised discriminatory practices, even though this awareness often coexisted with the perception that racism was ordinary or unavoidable, suggesting that recognising racism does not necessarily prevent its psychological effects.

Schools were the main setting in which participants experienced racism, although online environments also emerged as an increasingly important site of discrimination among young people. At school, adolescents described discriminatory attitudes and practices by teachers, unequal disciplinary practices, lack of safety, and social exclusion, illustrating how racism in everyday school life affected their mental health and willingness to study. These experiences are consistent with studies showing that school-based racism is associated with lower school connectedness, greater peer victimisation, and poorer mental health.^21^ However, our results suggest that the experience of racism at educational settings extends beyond discriminatory experiences to include unequal access to economic resources, further intensifying the mental and emotional burden.

Racism, both historical and ongoing, was seen by the participants as shaping and sustaining socioeconomic disadvantage, affecting adolescent mental health by producing material conditions that undermine wellbeing. This reflects the intersectional experience of being minoritised and living on a low income. Within these contexts, the internalisation of responsibility, sometimes described as individualised coping under structural disadvantage,^22^ can intensify stress, anxiety, and depressive symptoms. These pressures also shaped participants’ expectations for the future, corroborating evidence that structural constraints influence adolescents’ long-term outlooks in distinct ways.^23^ Extending previous research on interpersonal discrimination,^2^ this study shows that minoritised adolescents are aware not only of direct exclusion but also of the structural inequities that shape their everyday lives, and that navigating both personal discrimination and broader systemic constraints creates a distinct psychological burden.

Although participants primarily described the mental health consequences of racism, these experiences may also have implications for physical health across the life course. Previous research suggests that repeated exposure to discrimination during adolescence becomes biologically embedded through chronic activation of physiological stress-response systems, increasing allostatic load and altering cortisol regulation. These processes have been associated with shorter telomere length, epigenetic age acceleration, and increased risks of asthma, hypertension, cardiovascular disease, and other chronic conditions in adulthood.^7^ In this context, adolescents’ accounts of persistent injustice, insecurity, and psychological distress may represent not only immediate mental health consequences but also early experiences with the potential to shape long-term health trajectories.

Beyond its effects on mental health and potential implications for long-term health trajectories, racism also has more immediate effects on adolescents’ physical health. Rather than primarily attributing these outcomes to individual behaviours,^2^ participants emphasised the systemic pathways through which racism operates. These included inadequate nutrition linked to food insecurity and the broader consequences of economic hardship, and exposure to racialised violence, including physical aggression and violence justified or enacted by police and security actors. Poor living conditions, including insufficient sanitation and exposure to environmental pollutants, further illustrate how economic inequities compromise the physical health of minoritised adolescents. Environmental racism compounds these inequities by disproportionately situating minoritised communities in areas with higher levels of pollution and fewer resources,^24^ as experienced by Quilombola, Amantaní, Malaiyaha Tamil and Baganda participants.

### Adolescents’ Acknowledge and Recognition of Racism

It is important to note that our findings are based on adolescents’ perceptions, and racism is not always immediately recognised. While participants identified their experiences of racism, these experiences and their consequences were often minimised or normalised. This reflected broader societal tendencies to downplay structural inequality, as well as adolescents’ adaptive coping strategies for navigating these realities and wider contextual norms around disclosure, such as cultural differences in talking about emotions and expectations to preserve community harmony. Moreover, racism is often understood narrowly as a range of overt interpersonal aggressions explicitly referencing a person’s race, ethnicity, or migration status, which may limit the recognition of more subtle or structural forms of discrimination. The recognition of racism may be further complicated by seemingly ‘equal’ treatment, which maintains the status quo by failing to recognise that individuals and groups do not begin from the same social, economic, or historical position. For adolescents, navigating these dynamics can be particularly disorienting. When racism is minimised or disguised as equal treatment, its invisibility can create a kind of institutional “gaslighting,” invalidating young people’s experiences of marginalisation.

Several participants explained that they learned about racism over time and only then understood how to identify it. Youths’ capacity to identify and articulate racism develops over time and typically strengthens during adolescence as they are increasingly exposed to books, media, interpersonal conversations, and educational settings. This process is closely linked to the development of critical consciousness^25–27^ and is not solely an individual cognitive achievement but a socially mediated process shaped by cultural norms, educational environments, and the broader sociopolitical context.^28^ Attending to how adolescents come to recognise and name racisms globally is therefore central to interpreting their accounts and to understanding how developmental and social contexts shape both its recognition and its health impacts.

### Adolescent Voices and Partnerships for a Fairer and Healthier World

This research highlights adolescents’ priorities for action against racism, challenging adult-centric and deficit-based framings by positioning adolescents as knowledge-holders and agents of change. Across diverse countries, participants consistently identified priority areas for action, underscoring the coherence of their perspectives. For minoritised adolescents, meaningful agency extends beyond participation to include redistribution of power and resources that shape health, access to care, and broader determinants of wellbeing. The priorities identified by adolescents directly inform the implications for clinical practice and policy. These findings underscore the need to recognise racism as a structural determinant of adolescent health, shaping access to resources, opportunities, and supportive environments. They also support integrating sensitive enquiry about discrimination into routine mental health assessment and response. Health systems and service planners must ensure equitable access to culturally safe care by addressing barriers related to affordability, language, geography, and perceived differential treatment. The findings further call for coordinated policy action on social and environmental determinants of health, including housing, sanitation, and food access. This highlights the role of education systems and institutional practices in shaping long term health trajectories, indicating supporting policies that promote inclusion, fairness, and trust across sectors relevant to adolescent wellbeing.

### Limitations and Strengths

Spanning four continents and six countries, this study provides a rare global perspective on the forms of racism that adolescents face, while retaining the qualitative depth often lost in large-scale quantitative research. Shared methodologies supported cross-cultural comparison and analytic coherence, enabling identification of common patterns and local specificities. Iterative collaboration with in-country teams ensured culturally grounded interpretations, while partnerships with local organisations facilitated appropriate recruitment and interactions.

However, despite the benefits of vignettes for research on sensitive topics and facilitating examination of structural and institutional racism, participants’ responses may have been influenced by the method, reflected in the emphasis on educational settings. Although findings are presented at the country level, they present the perspectives of a single ethnoracial group in each site and do not capture the full diversity of minoritised groups within countries. The number of interviews and their durations varied across countries, though data saturation was achieved. Inviting young people to review the interview guide and share their demands moved beyond solely documenting their experiences towards a more action-oriented approach that acknowledges adolescent agency and anchors future advocacy efforts in youth voice, nevertheless, future research should aim to embed adolescent voices throughout the research process.

## CONCLUSION

This study contributes to understandings of racism and adolescent health by foregrounding how adolescents themselves perceive and experience racism, its impacts, and priorities for action.

Participants described racism as both interpersonal and structural, with consequences for exclusion, opportunity, and psychological well-being. Their accounts highlighted the psychological and emotional burden of navigating inequitable systems while being expected to overcome barriers through individual resilience alone. Addressing the effects of global racisms on adolescent health requires more than traditional health interventions. It calls for a health justice approach that tackles the structural drivers of inequity and prioritises equitable resource distribution, community participation, and inclusive policymaking to reduce health inequalities and create conditions that support both physical and psychological well-being. By recognising adolescents as key agents in identifying these inequities and articulating demands for change, this study highlights the importance of engaging adolescents in shaping policies and practices that advance social and health justice.

## Data Availability

The qualitative interview data generated and/or analysed during this study are not publicly available because sharing the interview transcripts could compromise the anonymity and confidentiality of participants. De-identified data may be available from the corresponding author upon reasonable request, subject to ethical and confidentiality considerations.

## AUTHORS AND CONTRIBUTORS

PdMS contributed to conceptualisation, study design, data collection, analysis, figure conceptualisation, and writing. SE contributed to study design, data collection, analysis, and writing. JM contributed to study design, interpretation of findings, and critical revision. SNíC, HC and SSK contributed to data collection, analysis, writing, and critical revision. MC, HPN, HM, AW, and KW contributed to data collection, analysis, and critical revision. AG and RL contributed to interpretation of findings and critical revision. MH contributed to figure conceptualization and design, and critical revision. ZB contributed to data collection and critical revision. CP contributed to critical revision. DD contributed to conceptualisation, interpretation of findings, and critical revision. All authors reviewed and approved the final manuscript.

## ACKNOWLEDGEMENTS

The authors would like to thank Blenda Milagros Abarca Díaz and Renan Espezua of HAMPI Consultores en Salud for their contributions to data collection; Touch the Heart Uganda and the community members of Kawempe District, Kampala, Uganda, for their participation and support of this study; Displaced CIC, Edmonton Community Partnership, and Indoamerican Refugee and Migrant Organisation for their support with participant recruitment; and the MHPSS.net team—Kaushi Jayawardena, Nandhini Devi Shankar Dass, Nithila Theivendran, Oenone Mills, Prapanchan Sornalingam, Sivatharsini Ravindran, and Vigitha Renganathan—for their assistance with facilitation, transcription, and translation.

