## Appendix for "Adolescent Voices on Global Racisms: Lived Experiences, Health Impacts and Demands Across Six Countries"

| Theme | Subtheme and description | Country | Quote |
|---|---|---|---|
| Perceptions of difference | Appearance: Differences in appearance related to skin tone, hair texture, facial traits | Brazil | So, being a Quilombola, being a woman and being black, I think there is already a very big difficulty, especially in high school, where you have that mockery: look at her hair, look at how she dresses, there is this whole issue. (Girl, Brazil) |
| | | Peru | Yes, I think they would be very different.The [non-minoritised character]'s parents would likely have more financial resources, and their appearances would reflect that. The [minoritised character]'s parents, on the other hand, would be farmers or artisans who live from tourism. (Girl, Peru) |
| | | South Korea | I also think she would look just like any other Korean. (Girl, South Korea) |
| | | Sri Lanka | Now Dharshi will wear all the new dresses, but Kala will wear what she wears at home or anything that her friends would have given her. This may be old clothes. She will stand out from what you see of children these days. (Boy, Sri Lanka) |
| | | Uganda | But for skin color, maybe Sam is lighter-skinned because some people with money tend to look like that. Dan might be darker because he's from a poorer area. (Boy, Uganda) |
| | | United Kingdom | Well there's definitely going to be some sort of appearance difference between Jamal and Oliver and considering the fact that they moved, he might have different characteristics to Oliver. He may have a different skin tone, different facial features, things like that. (Boy, United Kingdom) |
| | Cultural and religious background: Differences in traditions, customs, religion, habits, and clothing, as well as "culture" in the words of participants. Participants invoked the term *culture* often imprecisely or without clear definition. Participants' use of this language is itself socially mediated, shaped by the dominant discourses available to them. | Brazil | The Quilombola culture is different from the non-quilombolas, because they have their own identity. The quilombolas have a specific relationship with the land, with African dance, with cultural traditions. (Girl, Brazil) |
| | | Peru | The difference is that here we have other customs, in another place. (Boy, Peru) |
| | | South Korea | I'm not like them. I'm mixed race and they're native Korean, so I'm different, so I think it's hard for me to get close to my friends from high school because I think I'm different. (Girl, South Korea) |
| | | Sri Lanka | Since they have different ethnicities then culturally also they will be different. Even if Dharshi is culturally a Hindu, their way of celebrating would be different. - In cultures there will be rules that say 'the child should behave this way'. In Dharshi's culture these rules may not be there. In terms of culture, this child's [Kala's] freedom may be affected. (Boy, Sri Lanka) |
| | | Uganda | Some people I study with segregate me because I am Muslim, they think we are different. (Girl, Uganda) |
| | | United Kingdom | Because you adjust to like two different cultures. You're also trying to maintain your home culture, whereas- and also like the UK culture is for Layla. She will probably like- I don't know. it's just hard to |

| | | | |
|---|---|---|---|
| | We use *cultural background* not as a reductive category, but to distinguish shared practices and ways of being from markers such as appearance, language, or location. | | like- Sometimes it's just hard to satisfy both cultures because like with me, I know that my parents still are very concerned over the South Asian culture. They're like, "Oh, yes. You need to be doing this, this way because South Asian people do it this way." But I'm here. Like, I'm used to the UK culture now and then sometimes the parents don't like it when you mix the UK culture, because they don't agree with all the UK culture because they haven't been born in the UK and they don't know like, the reasons behind the culture either. (Girl, United Kingdom) |
| | Language: Difference in accent, languages spoken, or language proficiency | Brazil | Not applicable |
| | | Peru | Or the way they speak, that is, here we speak Quechua. And they don't understand them there, if they go to the city at all. Of course. It would be a form of discriminating. (Boy, Peru) |
| | | South Korea | First of all, she has become proficient in two cultures and languages from an early age due to the influence of multicultural background. (Girl, South Korea) |
| | | Sri Lanka | They may be from different ethnic groups. Also, they [Kala] speak the Malaiyaha language. They [Tharshi] speak English, so... (Boy, Sri Lanka) |
| | | Uganda | Not applicable |
| | | United Kingdom | It takes long to understand English and proper. Like, one thing's like learning English one and then the second thing is, like talking to young kids with English like, so you go to a class, then some of them will make fun of you for your English, and so you got to adjust to that maybe a year or something. (Boy, United Kingdom) |
| | Location: Place was considered a marker of race and ethnicity that participants linked to worse economic conditions, status and resources where minoritised adolescents have grown up and lived, such as in communities, plantations and ghettos | Brazil | Because here where I live, here is a community. It's like I'm telling you, Quilombola communities have a lot of fishing, there are people who fish, you know? And there is, there is Cachoeira, Cachoeira is the headquarters, it is the city, the community is the district of Cachoeira. (Boy, Brazil) |
| | | Peru | Yes, it's possible that Ana's parents are from the community and *Helen*'s parents are from somewhere else, so they'll look and act differently as well. (Girl, Peru) |
| | | South Korea | Not applicable |
| | | Sri Lanka | Because they (minoritised character) come from the plantation, and when they go to a big place, they point them out and think they are inferior, but since [non-minoritised character] comes from a good place, they give her first priority. (Girl, Sri Lanka) |
| | | Uganda | From my side, I think where they come from, one comes from a poor area while the other is from a rich area. (Girl, Uganda) |

| | | | |
|---|---|---|---|
| | | United Kingdom | In terms of I guess where they grew up, yes. I can tell that Layla, her parents don't speak great English so from that alone they're from different places. (Girl, United Kingdom) |
| | Economic resources: Differences in income and generational economic resources, included perceived expensiveness of clothing | Brazil | Her attitude, in a way, is very classic nowadays because, coming from a Quilombola community, a simpler community, sometimes we have difficulty socializing with people who we know have a better [economic] condition than us. (Girl, Brazil) |
| | | Peru | The [minoritised character] wouldn't have the same access too, because [minoritised character] doesn't have much money. And the [non-minoritised character] does, because if she has it from the city or elsewhere, she will buy [things]. Of course. (Boy, Peru) |
| | | South Korea | Having a multicultural background usually implies having a foreign parent. It's not easy for foreigners to earn a lot of money while working in a foreign land. So, that might make things harder financially. (Girl, South Korea) |
| | | Sri Lanka | Based on their appearance and their economical situation they won't be treated the same. (Boy, Sri Lanka) |
| | | Uganda | Yes, the main difference is where they live. Katanga is a slum, so [minoritised character] probably faces a lot of hardships with things like housing and food. Kololo is for rich people, so [non-minoritised character] has a better life. (Boy, Uganda)<br><br>But for skin color, maybe [non-minoritised character] is lighter-skinned because some people with money tend to look like that. [minoritised character] might be darker because he's from a poorer area. (Boy, Uganda) |
| | | United Kingdom | I'd say it's mostly about being poor, because if, for example, their parents were more wealthy, yes, even though they didn't speak great English, if they went, if they had private healthcare, typically in those places, you'd get respected well, by the way. (Boy, United Kingdom) |
| | Educational resources: Lower-quality education, limited course diversity (such as fewer language and arts options), restricted access to well-resourced schools, and reduced availability of technology for learning | Brazil | Not applicable |
| | | Peru | That is the difference, that one studies in large schools, as in Amantani there are no good schools (Boy, Peru) |
| | | South Korea | Not applicable |
| | | Sri Lanka | [non-minoritized character] will probably be going to a big school. That is why her teachers praise her a lot and encourage her. For [minoritized character] it's not like that. The teacher will put her down when she doesn't bring the books that she is told to bring. (Girl, Sri Lanka) |
| | | Uganda | In some homes they lack yet in the other everything is okay. Then education, there's one going to a good school and one in a bad one. (Girl, Uganda) |

| | | | |
|---|---|---|---|
| | | United Kingdom | Yes, not being born here, it might be quite hard for him, since if you're born here, you get used to the education system. So in here, it's not just about memorisation, but it's more about analysing, while back in my home country, it's more about just memorising stuff where I'm from, it's more about education, more about memorising stuff and self-analysing, which is quite difficult. (Boy, United Kingdom) |
| Recognition of racism | Minimized and normalized: Racism was conceived as something that "will always exist", leading to the feeling that "nothing can be done about it besides getting used to it" | Brazil | Not applicable |
| | | Peru | Not applicable |
| | | South Korea | So sometimes they joke around using stuff related to that, like with language and discrimination and stuff. But I know it's all a joke to them. And I mess around back, like maybe hit them once or twice, like in a playful way, so I don't really mind.<br>Researcher : Doesn't it still bother you, though?<br>Participant 11: At first, it kind of did. Like, my parents—if they heard stuff like that, they'd probably feel hurt. So in the beginning, I cared more. But now... not really. Because I know it's just joking between friends. (Boy, South Korea) |
| | | Sri Lanka | I mean... I have this thing where in anything I aim to understand it and act accordingly. I take it as it is by thinking 'This is how life is', 'That is how they will be ' and I won't let it bother me. I won't take it as a big thing and feel it. (Girl, Sri Lanka) |
| | | Uganda | Not applicable |
| | | United Kingdom | It's a bit hurtful, but then I've gotten used to it, like you just laugh about it now, because that's something that I feel like, every person of colour eventually is going to experience. (Girl, United Kingdom) |
| | Realisation and conscientiousness of racism: Awareness of naming racism started after experiencing, witnessing or hearing about it | Brazil | I think that, because he is a Quilombola, we are still living in a world that is racist, as much as they say no, the world is still racist. (Boy, Brazil) |
| | | Peru | I think racism is when a white, gringo tourist comes and they call us black, that here most of us are brown, of course. And the discriminations when they give you half a nickname or something like that, I don't know if they say it as jokes or... It makes you feel terrible. (Boy, Peru) |
| | | South Korea | He originally lived in Vietnam, so his Korean was a bit poor and awkward. And in elementary school, you know how if someone's like that, or if they mess up, like going poop at school or something and get caught, they get teased a lot for that kind of stuff. So he was teased like that back in elementary school. And even when we went up to middle school, because he had already been teased like that since elementary school, he didn't have any friends. (Boy, South Korea) |
| | | Sri Lanka | Because we come from an estate (a marginalized background), many people don't value us. And since we don't have a lot of money, people don't take us seriously. (Boy, Sri Lanka) |
| | | Uganda | yes, there's people in our society who don't like darker skinned people, yet we are all black. (Girl, Uganda) |

| | | | |
|---|---|---|---|
| | | United Kingdom | But it's like, oh, this is happening because of this. You like, understand type of thing. (Girl, United Kingdom) |
| | Internalization: Manifested as belief in and adoption of racist tropes and negative beliefs about their own minoritised group, leading adolescents to feel miserable and "not good enough" | Brazil | In the room where I study, in the school in general, some do not consider themselves, but most consider themselves quilombolas and work so that others who do not consider themselves, also consider themselves. Easily accept being a quilombola, being black, being a black woman, with black skin. They have this facility because the school coordination is always helping us with this, to self-identify. (Girl, Brazil) |
| | | Peru | Because those who have more money believe in themselves, they believe they believe in themselves. And those who do not have money allow themselves to be commanded by those who have money. (Boy, Peru) |
| | | South Korea | If the government only helps multicultural people, the general public might complain, saying, "Why do they get special treatment just because they're multicultural?" So, I think rather than providing specific benefits, the focus should be on raising awareness and promoting equality in opportunities. (Girl, South Korea) |
| | | Sri Lanka | Not applicable |
| | | Uganda | Not applicable |
| | | United Kingdom | So people can just base you off that and think you're dumb. And you can actually become dumb from that, like, you can get so low self-esteem, and from that you can actually, like, get lower grades, that actually happens. (Boy, United Kingdom) |
| Forms and contexts of discrimination | Lack of access to resources and infrastructure: Included less access to food, sanitation, technology, transportation, spaces and facilities for exercise, clothes, and leisure options | Brazil | Because, like, many times, when there is no way for the bus to pass because there is no paved road and it has rained a lot. The people in the bus get off and have to walk the rest of the way, you know? And it is a path with a lot of mud. Then, many times, in order not to walk barefoot on this path, they wear shoes. When you get to school, the shoes are very muddy, you get the school dirty, you know? And other people, seeing that, the dirty shoes, start making fun of this person's face, start talking badly (behind his back), you know? (Boy, Brazil) |
| | | Peru | The [non-minoritised character] could eat richer things, like chicken or meat, while the [minoritised character] would only eat what is produced on the island, like goose or potato. (Girl, Peru) |
| | | South Korea | Not applicable |
| | | Sri Lanka | If there was a shop in the plantation, women could buy pads during their periods, but many people only use cloth. (Girl, Sri Lanka)<br><br>Maybe they wouldn't even have a proper room to sleep or study. Water facilities… Maybe some don't even have electricity. Could be like that. Maybe they don't have a proper kitchen to cook in. Even the toilets might be in poor condition. Something like that. (Girl, Sri Lanka) |

| | | | |
|---|---|---|---|
| | | Uganda | The food from one end is good and on one end bad, same with the uniforms for school, everything is different because of where they come from. (Girl, Uganda) |
| | | United Kingdom | I feel like there will be a difference in the food, clothes and medicine Jamal would gain compared to Oliver. Even though like, I say like, as I say, with like food and stuff like that, compared to Oliver's parents, it won't be as drastic a difference compared to like medicine and clothes. (Boy, United Kingdom) |
| | Worse future opportunities and conditions: Included discrimination during job searches, lack of parental support due to long working hours, limited local employment options, lack of citizenship, language barriers and the knock-on effects of poorer educational experiences and constrained educational opportunities | Brazil | We go through great difficulty even to find a job, the color of our skin, our hair, the way he talks, the way he walks, his clothes. I think all of this encompasses a more difficult life. (Boy, Brazil) |
| | | Peru | [minoritised character] may be worried about her future. She will wonder what she is going to do after she finishes school, how she is going to continue her studies if she wants to be a doctor. Those thoughts may affect your focus in school. (Girl, Peru) |
| | | South Korea | Not applicable |
| | | Sri Lanka | Even for jobs – compared to Tamils, it's the Sinhalese who mostly get government jobs. For Tamils, jobs mean working in shops and such. They treat us in a lower way. That's the kind of work Tamils are taken for. (Girl, Sri Lanka) |
| | | Uganda | Researcher: Does it affect the opportunities [minoritised character] might get? How? Why do you think that?<br>Participant: Yes because When you go to ask for a job, they first look at where you studied, primary, secondary, university. And you can't compare a person who comes from one school to another. When a person goes from a good school they say yeah he went to a good school even though the one from a bad school won in performance. (Girl, Uganda) |
| | | United Kingdom | Kind of yes, because if you're a migrant, it's more hard for you to earn money, in the case that since you enter the system so late, like the job system so late, it's quite hard for you to earn more money than people who live there since their childhood. (Boy, United Kingdom) |
| | Worse treatment in educational institutions: Was mainly from peers through bullying or isolation. Teachers also discriminated, through expecting less from minoritised adolescents or | Brazil | So, being a quilombola, being a woman and being black, I think there is already a very big difficulty, especially in high school, where you have that mockery: look at her hair, look at how she dresses, there is this whole issue. (Girl, Brazil) |
| | | Peru | Participant: No, they discriminated against me and my friend.<br>Interviewer: Another colleague?<br>Participant: Yes.<br>Interviewer: And did it affect you?<br>Participant: yes, a lot, because...<br>Interviewer: Sure...<br>Participant: I feel bad when they say ugly things to me. (Girl, Peru) |

| | | | |
|---|---|---|---|
| | through assumptions of wrongdoing. Being treated poorly discouraged adolescents from staying in school | South Korea | Researcher: Do you think Jina has experienced discrimination due to her background, either at the hospital or online?<br>Participant: I don't think so. (Boy, South Korea) |
| | | Sri Lanka | In some schools where there is Tamil medium and Sinhala medium, the priority is given for Sinhala medium. They will try and put them only first for everything. The Tamil medium won't even have an idea that something like this is there. We only know when they come and tell after other students go and attend things like that. Things like that shouldn't be there. (Girl, Sri Lanka) |
| | | Uganda | Some people I study with segregate me because I am Muslim, they think we are different. (Girl, Uganda) |
| | | United Kingdom | There was a time we were in maths class and this white person, this white guy, he was saying probably the most racist thing I've ever heard on this earth. He was saying everything under the sun and yet the teacher didn't say a single thing to this boy. (Girl, United Kingdom) |
| | Lack of availability and accessibility to good education: In segregated settings, schools attended by minoritised adolescents were often poorly resourced and lower in quality compared to those in more privileged areas. Challenges included long distances, lack of transportation or safe routes, limited language support or translation in multi-ethnic contexts, and unequal access to opportunities within the school and in school-based competitions and events | Brazil | Because we are not the majority, we are the minority in everything. We don't go on a large scale to other universities. Practically, in large universities, if you meet two quilombola people, I think it's a lot. (Boy, Brazil) |
| | | Peru | Because [the non-minoritised character] has a more modern school and that serves better. (Girl, Peru) |
| | | South Korea | Not applicable |
| | | Sri Lanka | Yes, it did. For example, our music teacher left, and even though they kept saying again and again that they'll bring someone, it took a 1 month to 1.5 months before a new teacher was assigned. During that time, the students learning music found it very difficult—even to attend the class. Even now, in our school, there is no teacher for the Health subject. Even if we raise it, nothing is really considered. Another teacher from a different subject is asked to teach it. Whatever the case, how effective is it really for us to learn from someone who is not trained in that subject? That's something to think about. (Girl, Sri Lanka)<br><br>Hmm… hmm… the facilities might be limited. Because they may not go out much for external things [opportunities]. So going for higher studies might not be common. In the estate, half the people say, "With so little, why study so much?" There's a chance people might say things like, "Better to go for work halfway through." Some parents might let them go to work. Others might say no, the child must study. (Girl, Sri Lanka) |
| | | Uganda | He will always be panicking, he lives in a bad place and even the school isn't good. (Boy, Uganda)<br><br>In education, like paying fees, chasing him for late payment, and also when it comes to senior 6 (final secondary school year) it's expensive and requires many things and he might not afford. (Girl, Uganda) |
| | | United Kingdom | Some of them can just like, because the English, they can just like, get low grades. (Boy, United Kingdom) |

| | | | |
|---|---|---|---|
| | Lack of availability and accessibility to healthcare and medicine: Included lack of staff, language difficulties, insufficient income to afford private care, complex bureaucratic requirements for immigrants, long distances to health centres, medication shortages, and overcrowded facilities | Brazil | It's limited access, distance can also affect health a lot. Because, for example, a person from a community that is far from the health center here, for example, here in my community there is a health center, but there are other communities that are a little far away. Then there is someone feeling sick. This person, (to call the ambulance, the ambulance arrives to refer to the health center here. If there is no way to solve it at the health center, it has to be referred to another place that can solve it. And I think the distance doesn't help much). (Boy, Brazil) |
| | | Peru | Because [the minoritised character] does not have the same economic resources and is from Amantaní, so she cannot access better services. (Girl, Peru) |
| | | South Korea | Not applicable |
| | | Sri Lanka | Now they [Tharshi] are staying in a place like a town, right? So, because they are in a town-like place, the hospital will be nearby. So, if there's any emergency, they'll go and come back easily. But in the hill country (estate sector -'malaiyagam' is the term used to describe it), to go from the hill country to the town is a bit difficult. There aren't many vehicles there either. One must bring a vehicle themselves and go. In that time, their health might be affected. (Boy, Sri Lanka) |
| | | Uganda | Medicine, when they go to the clinic to buy, and he tells the person there that he doesn't have enough money , the person will give him a cheaper substitute and sometimes this can't treat him well. (Girl, Uganda) |
| | | United Kingdom | I know people, not in my case, because my dad knows a lot, really good English, but I know a few people whose parents are like, not that good like they can't really speak that much, or they need assistance. It would usually be my friends, so their child translating for them. Yes, so where, even if they come along with them, they would usually be the ones, the kids would usually be the ones that are translating for their parents. (Boy, United Kingdom) |
| | Worse treatment in health institutions: Participants or their family members experienced neglect, dismissal of their symptoms and concerns, and outright discrimination in health institutions | Brazil | It would depend on whether the doctor was a quilombola, because if he were a quilombola he would treat him the same way, but if he wasn't, he would treat him unequally. Because there are many doctors like that. (Girl, Brazil) |
| | | Peru | They would do it in a serious way. For example, they might say to [the minoritised character], "You can't come in now, come later." But if someone with more resources arrives, like [the non-minoritised character], they attend to her immediately and with more courtesy. (Girl, Peru) |
| | | South Korea | I have a foreign registration card, but I only had a photo of it saved on my phone. I showed it to them at the new clinic, but they said it wouldn't work and asked, "Are you a foreigner?" When I said yes, there was this awkward silence—maybe just 2 or 3 seconds, but I definitely noticed it. (Girl, South Korea) |
| | | Sri Lanka | Hmm… hmm… there are a lot of Tamil and Sinhala children at the hospital. In that case… if you look at Tharshi, she'll go to a big hospital. She can pay money and go get seen. But for [minoritised character]… [minoritised character] will go to the government hospital. There are more chances of Sinhalese people being there. So, because [minoritised character] is Tamil, there will be discrimination based on ethnicity. |

| | | | |
|---|---|---|---|
| | | | (Girl, Sri Lanka) |
| | | Uganda | Res: Yes, I've felt it before. One time, at the health center, I waited for hours, but people from the majority religion in Kawempe were treated first. It felt like I didn't matter just because I'm Christian. (Girl, Uganda) |
| | | United Kingdom | But literally the only person that I hold those grudges against is the so-called NHS community or the doctors that I dealt with at the hospital in London because you're assuming, based off of my skin colour that I'm exaggerating because I heard one of the conversations that they had with the nurse. That's why I'm saying exaggerating because once I left the hospital room a little bit late, I heard the doctor speak with the nurse, they're just exaggerating, just give them paracetamol, antibiotics and what not. (Boy, United Kingdom) |
| | Verbal and written insults and attacks: Occurred on social media or in person by friends and strangers in public spaces. Included overt and covert racist "jokes" and name-calling | Brazil | Mean comments. To say: "You black, monkey", these things. (Boy, Brazil) |
| | | Peru | I think that someone from the countryside would be criticized, criticized on TikTok or on any social network. (Boy, Peru) |
| | | South Korea | I have a friend who is half Cambodian. He is a Korean who grew up in Korea, but sometimes friends call him "negro". (Boy, South Korea)<br><br>Participant: During the Vietnam War, when Korean soldiers went to Vietnam and had children with Vietnamese people, they called those kids "Lai Dai Han." So they tease me with that.<br>Researcher : How do they tease you with that?<br>Participant: They just call me "Lai Dai Han." And in our current math unit, there's this symbol called a radian, right? They use that too and call me "Radian," just messing around. And since "Lai Dai Han" kind of sounds like "Ratatouille," they sometimes call me "Rata" or something like that. (Boy, South Korea) |
| | | Sri Lanka | It's like… sometimes people don't even respond when she talks or asks for help. They ignore her needs and scold her instead. Sometimes they say things that emotionally hurt her. (Girl, 16, Sri Lanka)<br><br>They see it like that, don't they? Some people say things like, "They're upper caste, lower caste—we shouldn't mix with them." That kind of talk. (Girl, Sri Lanka) |
| | | Uganda | Not applicable |
| | | United Kingdom | Yes, I feel like, especially with like, apps such as like Instagram compared to like TikTok, Snapchat and stuff like that, I feel like with Instagram, like the comment sections are like more like honest and brutal and stuff like that. So I feel like with like apps such as that, where the comment sections are open, I feel like the comments that Jamal would receive, like being that open and posting about stuff like that, I feel like the audience would like be less accepting of him in a certain way and be more like questioning his like, what he does on social media compared to someone like Oliver, in a sense. (Boy, United Kingdom) |

| | | | |
|---|---|---|---|
| | Discriminatory gazes: Included nonverbal expressions of judgment, exclusion, or condescension in the form of looks and stares | Brazil | In things, I can say that Quilombola people attract looks, it may not be about compliments, but it also has a lot of judgments. (Girl, Brazil) |
| | | Peru | No, they would look at him ugly, saying: Serrano, Serrano. (Boy, Peru) |
| | | South Korea | Not applicable |
| | | Sri Lanka | But in [minoritised character]'s case, people often look at her with disdain, treat her differently, and lead her away in another direction. (Girl, Sri Lanka) |
| | | Uganda | No, they wouldn't be treated the same. Dan might be ignored or looked down on because he's from a slum. Sam would probably get more respect because people assume he has money. (Boy, Uganda) |
| | | United Kingdom | But at the same time, it's like some of the people in general, outside, sitting the waiting room, they're just looking. And that's everyone, that's, that's what everybody does. They can look you. (Boy, United Kingdom) |
| | Assumptions of wrongdoing and unequal disciplinary measures: Included profiling and harsher disciplinary measures compared to non-minoritised peers by security guards, police and teachers | Brazil | At the bank, recurrently as it is currently happening in the newspapers, there are always blacks who end up being murdered, end up being arrested for the fact that they are black. And many times they are innocent people and end up taking the chicken jumping and don't even know why. (Boy, Brazil) |
| | | Peru | Not applicable |
| | | South Korea | Even my homeroom teacher didn't seem to care why I wasn't coming to school. They didn't ask why—I think it was more like, "Why aren't you coming? You should come to study; you're in the third year now, and it's important." I already know that, but no one asked me why I wasn't coming to school. (Girl, South Korea) |
| | | Sri Lanka | One time, a Sinhala boy brought drugs to school. An uncle [frequently used as a polite form of address for older men, regardless of whether they are actually related to the speaker] on the road saw and informed the school, and also reported it to the police. But that issue never spread within the school. The boy was suspended for a week, and then he was back in school like normal. On the other hand, a Tamil boy once brought juice and chocolates to school to celebrate his birthday. That was blown out of proportion, and people spread rumors that he was bringing drugs. The entire school heard about it. This kind of discrimination exists in our school. (Boy, Sri Lanka) |
| | | Uganda | Not applicable |
| | | United Kingdom | I would say like walking into like certain stores, especially like the outskirts of London, where it's not as diverse, like people like watch you more closely, or they think you're up to no good or something like that. And I think I remember one time going to a Sainsbury's, I think I walked past the fruit section, and I was looking, because I didn't buy anything from the fruit section, so I got what I wanted from another section. So I was coming out, and the security guard, he actually saw me go through the fruit section, and he thought I put something in my bag. And he was like, "Oh, where's the fruits? Where's the fruits?" So I |

| | | | |
|---|---|---|---|
| | | | was just confused. I said, “I don't have any fruits with me.” He told me to open my bag. (Boy, United Kingdom) |
| Health consequences of racism | Emotional wellbeing and mental health: Included poor mental health broadly, ranging in specific cases from anger to sadness and possibly depression. In rare instances, participants also mentioned suicidal thoughts. | Brazil | Because depending on the quilombo he lives in, he may not have many medical conditions, these things that can be... because it is difficult for him to live with the differences that are placed between him and other colleagues or even his mental health can end up going down the drain, because of the difficulty he has to relate to other colleagues. (Boy, Brazil) |
| | | Peru | But there are also people who do not treat everyone equally, and they bully, which can affect them emotionally. (Girl, Peru) |
| | | South Korea | Researcher 1: Right. How do you think such cultural factors influence Gina’s health, whether mentally or physically?<br>Participant: Honestly, I think they do. Even though my parents are Vietnamese, someone else’s prejudice against Vietnamese people could lead to discrimination. These issues might result in mental harm. Since I understand how these students feel, I think that’s possible. (Boy, South Korea) |
| | | Sri Lanka | Our feelings… We feel a bit angry and sad. We wonder, “Why are they doing this, just to us?” That kind of feeling arises. (Girl, Sri Lanka) |
| | | Uganda | He will always be panicking, he lives in a bad place and even the school isn’t good. (Boy, Uganda) |
| | | United Kingdom | I mean it could decrease my mental health, make me feel like I'm not worth going into these good universities, get a good education and stuff. (Girl, United Kingdom) |
| | Physical impacts: Included physical injuries due to racial violence, loss of appetite and generalised illness due to poor mental health, lack of nutritious food and exposure to pollution and dirtiness | Brazil | And in my case, it affects because I wake up at 3 am to be able to do this 40-minute walk, more or less as well. This walk is even good, but at the same time it is bad because I still take coldness. But when I return home, I take the 5-year-old boat, go to school and also arrive very early at school. I also don't eat properly. It affects both, the physical and the mental. And even when I return home, it's an hour and sometimes I can't find the boat and I have to stay until 3 pm to be able to get home and have lunch. When I arrive, the hunger has already died in the body and sometimes I am not so hungry and then it is harming me, both mentally and physically. (Girl, Brazil) |
| | | Peru | Not applicable |
| | | South Korea | If someone’s mental health declines, they may lose their appetite, stop going outside, and eventually, their physical health could deteriorate too. (Girl, South Korea) |
| | | Sri Lanka | Since they are economically lower, they will only be able to get things for the house according to the money they have in hand. This can be less healthy. They won’t be in a situation to buy things that are healthy. So the food they cook at home can be less health. (Boy, Sri Lanka)<br><br>If they don't eat properly, they get gastritis, if there is a lot of pressure then that can lead to a heart attack. (Girl, Sri Lanka) |

| | | | |
|---|---|---|---|
| | | Uganda | Poor sanitation causes diseases and this means they won't be able to go to school. (Girl, Uganda) |
| | | United Kingdom | I feel like, if it's like, really bad, it can trigger something physically like, it can be mentally at first, and after that it comes to be like physical harm. (Girl, United Kingdom) |
| Buffers and protective mechanisms | From family and friends: Through advice, friendship, comfort, opportunities for reflection, distractions from worries, and protection from bullies. Family was perceived as a place of constant support, independently of changes in life | Brazil | My mother thought I was becoming depressed, so she even paid for me to see a psychologist to help improve my self-esteem. She started talking with me a lot about these things. Sometimes I had anxiety attacks, and my mother would talk to me and call me beautiful, beautiful. She would tell everyone who came close to me, even family members, about it so they could help build my self-esteem. My mother helped me a lot with that. She would say: 'Look, that girl says she's ugly, but she isn't ugly. People say that about others too, but it isn't true.' The boys used to say she was ugly, that her hair was too long or too full, and sometimes they also said her hairstyles were ugly. What really helped me was my mother encouraging me and building my self-esteem through these conversations. I also went to the psychologist. Not for long, but I did go. She shared a little of her own story as well, saying that she had suffered too but that it didn't define her. She said you have to move forward and work towards your goals, to become a better person. Not better than anyone else, but a more educated person and someone with a profession. She didn't want me to be without a profession like my mother and father, who don't have formal professions but work as fishers and shellfish gatherers. (Girl, Brazil) |
| | | Peru | I had a friend, and he recommended me: Don't listen to people, instead focus on studying, on books. (Boy, Peru) |
| | | South Korea | I think having conversations would help. From my experience, though I haven't lived that long, I've found that communication and talking with friends are really important. When I talk with someone, sometimes my worries feel lighter without me even realizing it. (Girl, South Korea) |
| | | Sri Lanka | amil people don't see any difference and they treat everyone equally. There is no unity in the way others behave. Seeing this makes me feel worried like Why am I still a slave...Makes me feel worried. When I feel like this I talk to my friends. (Boy, Sri Lanka) |
| | | Uganda | To feel better, I think he can sit down and tell his mother about how he feels. (Girl, Uganda) |
| | | United Kingdom | I feel like, if she can have a conversation with her family and be like, "Oh, I feel like this, and this, this." Then her family might move her to somewhere that's more diverse, with different people of different races, I feel that will help. But like, If it's something in the moment, like, oh, this just happened to me right now. What should I do now? She probably goes to a trusted adult and be like, "Oh, this happened to me." This, this, that and explain how she feels about that and how she wants that to, like, change. (Girl, United Kingdom) |
| | Within communities and local environment: Through a sense of protection from | Brazil | So, most quilombola communities come with agricultural products, they don't come with anything exported, nothing canned, you know? And I think that this diet can make it healthier. (Boy, Brazil) |
| | | Peru | I don't think so. Being Quechua can be an advantage, for example, if one day [the minoritised character] becomes a doctor and has to attend to patients who only speak Quechua. If you know the language, you will be able to communicate with them better. (Girl, Peru) |

| | | | |
|---|---|---|---|
| | discrimination within local areas, and of strength and pride in one's communal identity | South Korea | Participant 11: In my case, my mom speaks both Vietnamese and Korean, and sometimes she helps people who visit her workplace and struggle with Korean, like at the police station. She translates for them from Vietnamese to Korean and vice versa.<br>Researcher : How do you feel when you see your mother playing this bridging role?<br>Participant: I respect her a lot. Moving from Vietnam to Korea wasn't easy, and she worked hard, studying while taking care of us. She even got her driver's license and now drives well. Seeing her do all this makes me admire her. (Boy, South Korea)<br><br>Researcher : Do you also see your mother's background as an asset?<br>Participant: Yes, language itself is a great asset. Since Vietnamese has a word order similar to English, it makes learning English easier. Chinese has a different writing system, which can be hard for exams, but speaking it isn't as difficult because I already use tones in Vietnamese. (Boy, South Korea) |
| | | Sri Lanka | All this time they were thinking about us differently, about the Tamil people. After we went and they got to know us, how we are, and the bonding between them and our religion. The feeling of 'How we can work together as one' was something that came to them. Because of that they have as much as possible bonded with us. Now in my village, there is a good impression of Tamil people now. We have as much as we can to help build that. (Boy, Sri Lanka) |
| | | Uganda | Not applicable |
| | | United Kingdom | …I, when it happened, I think I started to surround myself with Latin people, so…which made me feel, honestly, much better, I felt like I had already found people with…where…where I could feel good and so…so I think that made me feel better, surrounding myself with the people I wanted and…and nothing, that. (Boy, United Kingdom) |
| | Within institutions and systems: In the educational system, through affirmative actions for minoritised students to access education, anti-racist curriculum at schools, trained school staff, and cultural adequacy of served foods. In the health system, through diversity in staff, and understanding of language and cultural needs | Brazil | I learned it in high school. In addition to being a black school, 90 percent of the students at the school where I study are black. Here at my school, the students are from Quilombola communities . Because the school where I study serves 18 communities in Cachoeira. In addition to being a black school, they teach there, the teachers focus a lot on these things: discrimination, racism… And there are always lectures, classes, you know? and even though we are subject to living this today. I think that as time goes by, we learn. (Boy, Brazil) |
| | | Peru | Here at my school we have a psychologist, but not all students visit him because of shyness. Those who do get good results and improve emotionally. It would be good to encourage more use of these resources. (Girl, Peru) |
| | | South Korea | There's also an affirmative action programme for multicultural background families, so it could make things easier for her to some extent. But this affirmative action programme is grouped together with others—like for children from large families or families of national merit—so it's not that advantageous, but I still think it might have some impact to a certain extent. (Girl, South Korea) |
| | | Sri Lanka | So some teachers wouldn't let them (the students) be sent (to do the work), they would hold them back. 'They shouldn't go, they should focus on studies.' At those times, we would feel happy. That they were |

| | | | |
|---|---|---|---|
| | | | being supportive. (Girl, Sri Lanka) |
| | | Uganda | Not applicable |
| | | United Kingdom | Not personally, but they do every year, they remind us of, you know, certain guidelines and certain rules. (Boy, United Kingdom) |
| Demands to make the world a fairer place | Equal treatment: This was the most cited demand, referring to the desire not to be discriminated against and grounded in the belief that "everyone is equal" | Brazil | I think it is important to stop looking at judgment, to appear more opportunities and not to commit more acts of racism. (Boy, Brazil) |
| | | Peru | Yes, that we should all be treated equally. (Girl, Peru) |
| | | South Korea | Researcher: You mentioned earlier that multicultural families often face financial difficulties. Are there any policies or societal measures you think could help address this?<br>Participant: Changing societal perceptions is key. There's often a sense of superiority in our society toward certain ethnic groups like Southeast Asians or Chinese. Breaking that mindset and recognizing them as equal members of society is crucial. (Boy, South Korea) |
| | | Sri Lanka | We—ah, friends—I think all of us should just get along equally. Like, just because someone is from the estate, thinking "he shouldn't be my friend" or "he's from the town, so he's okay"—those thoughts shouldn't exist. We should treat everyone equally and just be friends. (Boy, Sri Lanka) |
| | | Uganda | I think no one should be discriminated at all (Girl, Uganda) |
| | | United Kingdom | Then of course it's people who are on the streets, family, friends, people related to you, people you know and shop owners and things who really need to raise light towards treating everyone equally because the government can't really force you to treat everyone equally. (Boy, United Kingdom) |
| | Support from family and friends: With parents and families playing an important role in establishing values of equality and non-discrimination, as well as family and friends providing support and standing up to racism together | Brazil | I think that the young person, that first he should have the support of his family, his mother, his father. Then, the people who talk, be his true friend, encourage him to do the right thing. Do not do anything wrong. And then, he believes in himself and moves on with life. (Girl, Brazil) |
| | | Peru | The family can also help, supporting their children and understanding them. We, as children, can help our parents understand what is right or wrong, especially if they did not have the opportunity to study. (Girl, Peru) |
| | | South Korea | I think it would help if she had more people to lean on—teachers, close friends, or even just a more supportive environment. (Boy, South Korea) |
| | | Sri Lanka | The mother and father's support for the child and the surrounding context. We also should create an environment that helps them to face challenges. Parents should make the children aware earlier that these types of problems may arise. This will help the child be more prepared, and not respond in fear, and will be able to focus on their work. There should be someone in the village or country who can tell them the right and wrong of life situations. (Girl, Sri Lanka) |

| | | | |
|---|---|---|---|
| | | Uganda | Me and my friends should love each other. (Boy, Uganda) |
| | | United Kingdom | Yes, he could surround himself with a social circle where he feels better or he could improve the way he sees what happens around him, like the teasing and that, he could not give it importance and so on. (Boy, United Kingdom) |
| | Anti-racist education: With anti-racist education in schools, through cultural education and racism awareness | Brazil | I think that all Quilombola schools have to show students that everyone is equal, regardless of race, color, gender, group, community. I think all schools must show that everyone is equal. I think the school has to address this, because it is within the school itself that we see that. (Boy, Brazil) |
| | | Peru | Teachers should talk to students to tell them that they shouldn't discriminate. (Girl, Peru) |
| | | South Korea | I think focusing on those educational activities is important. While some students complain, “Why are we doing this after exams?” I think we should recognize their value and pay attention. (Girl, South Korea) |
| | | Sri Lanka | To avoid that kind of situation, we have to teach children right from when they’re small—not to look at caste or religion. If we raise them with that mindset from a young age, when they go out into society, they’ll feel a sense of unity. They won’t think in those divisive ways. It’s only because the elders taught them these things that the younger ones start thinking like that. (Girl, Sri Lanka) |
| | | Uganda | We need to teach children about love and that we are all equal. Schools and churches should work together to promote unity and understanding. (Boy, Uganda) |
| | | United Kingdom | So obviously I understand even if in a great school you’re learning about things that some people will have their fixed mindset but the fact that people aren’t trying to teach people of diversity, of different things, culture, different religion, difference in the way that we look. That in itself is an effort that we should be doing and it’s clearly lacking. (Girl, United Kingdom) |
| | Better education and job opportunities: As a means for minoritised adolescents to have better lives and futures, particularly to escape from structurally determined circumstances | Brazil | The increase in vacancies for universities. Better education in the communities and an additional investment by both the State and the municipality in the education of the community. (Boy, Brazil) |
| | | Peru | Through education, we are educating ourselves through school. (Girl, Peru) |
| | | South Korea | I’m not sure about elementary schools, but in middle school, there was this one kid that I mentioned earlier who was teased in the elementary school. If a teacher had intervened and told students to be more accepting, it could have helped. In our school, we also had Russian students. They didn’t seem to experience discrimination because they made friends quickly. If teachers encouraged students to include foreign students and helped them integrate, maybe that one teased student wouldn’t have had such a hard time. (Boy, South Korea) |
| | | Sri Lanka | For any job, there should be good opportunities for Tamil people too. If they don’t, and if they separate us, our chances get reduced. Then some people will continue to discriminate. (Boy, Sri Lanka) |
| | | Uganda | They could offer scholarships to children from poorer families who perform well. (Girl, Uganda) |

| | | | |
|---|---|---|---|
| | | United Kingdom | Education a hundred percent, fix that. (Girl, United Kingdom) |
| | Rules and repercussions: Needed to inhibit racism and be educational to the discriminating person and others. Teachers and directors play an important role in assuring this at schools; however, participants felt that consequences of discrimination and racism were poorly enforced | Brazil | Raise the flag of “no to racism” and punish more rigorously people who discriminate against citizens. (Girl, Brazil) |
| | | Peru | Expel students who do that. (Boy, Peru) |
| | | South Korea | The government should improve working conditions for foreign laborers. I’ve seen news reports of foreign workers not getting paid properly. The government should enforce stricter punishments for such cases and improve their treatment. (Boy, South Korea) |
| | | Sri Lanka | Not applicable |
| | | Uganda | Not applicable |
| | | United Kingdom | Yes, we have some policies about it, they have extra rules to protect people who have different identity, identities, you know, be more strict to people who pick on them. (Boy, United Kingdom) |
